# Overdiagnosis and treatment of COPD in nonagenarians

**DOI:** 10.1101/2022.01.21.22269644

**Authors:** Mateo Tole, Christian J. Ascoli, Min Joo, Israel Rubinstein

## Abstract

**Background:** The prevalence of COPD is increasing with age. However, the effects of age-dependent decline in lung function on diagnosis and treatment of COPD in nonagenarians are uncertain.

**Objectives:** To determine performance of spirometry, prescription of COPD medications, and COPD-related acute care visits and hospitalizations in patients 90 years and older with physician-diagnosed COPD.

**Methods:** Health records of 166 consecutive patients 90 years and older with physician-diagnosed COPD at a university-affiliated medical center in Chicago were reviewed. Pertinent demographic, clinical, and physiological data were extracted.

**Results:** Patients were predominantly ex-smoker (96%), African American (52%) males (96%). Sixty patients (36%) had no spirometry testing on record. Of the remaining 106 patients, 11 (10%) had baseline FEV_1_/FVC≥0.70, 24 (23%) had post-bronchodilator FEV_1_/FVC ≥0.70, 28 (26%) had FEV_1_/FVC <0.70 and ≥LLN, and 43 (41%) had FEV_1_/FVC <0.70 and <LLN. Thus, only 71 of 166 patients 90 years and older (43%) fulfilled the Global Initiative for Chronic Obstructive Lung Disease (GOLD) recommendations. Nonetheless, COPD medications, predominantly short-acting β_2_ agonists and long-acting muscarinic antagonists, were prescribed to 95 of the 166 patients (57%). No significant differences in prevalence of co-morbidities and prescribed COPD medications, including systemic corticosteroids and anti-infectives prescribed during unscheduled healthcare visits and hospitalizations, were found between the four groups.

**Conclusions:** These data suggest that a large proportion of nonagenarians at our medical center are overdiagnosed with and treated for COPD. A larger, multi-center, prospective study is warranted to support or refute these retrospective observations.

## Introduction

The U.S. nonagenarian and older population now comprises 4.7% and is estimated to reach 10% by 2050 [1]. This trajectory is important because the incidence of COPD is increasing with age due to increased longevity and widespread tobacco smoking in the 20^th^ century [2, 3]. Despite a large body of medical literature, however, the effects of age-dependent decline in lung function on the diagnosis and treatment of COPD in nonagenarians are uncertain [4-12]. Conceivably, overdiagnosis of COPD in this age group could be associated with serious adverse events due to prescribed COPD medications, including administration of systemic corticosteroids during presumed acute exacerbations of COPD [11, 13-16].

Spirometry testing with forced expiratory volume in one second over forced vital capacity (FEV_1_/FVC)<0.70 after inhaling a short-acting β_2_-agonist is the mainstay for the diagnosis of COPD [12]. However, FEV_1_ declines more rapidly than FVC with aging leading to age-dependent decline in FEV_1_/FVC in healthy, non-smoking, elderly individuals [4-10, 16]. To account for this decline, a variable FEV_1_/FVC threshold derived from age-adjusted lower limit of normal (LLN) was developed to diagnose COPD [10, 17-20]. However, post-bronchodilator FEV_1_/FVC <LLN is not considered by the Global Initiative for Chronic Obstructive Lung Disease (GOLD) guidelines in diagnosing COPD among nonagenarians [21].

To begin to address this knowledge gap and unmet medical need, the purpose of this retrospective study was to determine performance of spirometry, prescription of COPD medications, and COPD-related acute care visits and hospitalizations in patients 90 years and older with physician-diagnosed COPD.

## Materials and Methods

Electronic health records of patients 90 years and older with physician-diagnosed COPD (ICD-10 codes: J44.0, J44.1, and J44.9) at the Jesse Brown VA Medical Center (JBVAMC), were reviewed on August 5, 2016, and July 28, 2018. Forty five of 211 patients thus identified were excluded because no documentation of COPD diagnosis was reported in physician notes.

Demographic information, smoking history, pertinent co-morbidities, spirometry testing, and COPD medications were abstracted from the remaining 166 health records. In addition, visits to urgent care and emergency department and hospitalizations for respiratory symptoms during the year preceding health record review were retrieved. Prescriptions of oral or intravenous corticosteroids and anti-infectives during urgent care and emergency department visits were tabulated.

Spirometry data were accepted only if pre- and post-bronchodilator flow-volume loops were adequately performed based on the American Thoracic Society/European Respiratory Society guidelines [22]. In each case, the most recent available spirometry report was reviewed. Patients with adequate spirometry were classified into 3 groups based on post-bronchodilator FEV_1_/FVC: 1) FEV_1_/FVC≥0.70; 2) FEV_1_/FVC<0.70 and >LLN; and 3) FEV_1_/FVC<0.70 and <LLN (10, 17-19). Lower limit of FEV_1_/FVC values for men and women were set at 0.627 and 0.635, respectively, based on NHANES III (20).

### Data and statistical analyses

Data are presented as means ± standard deviation where appropriate. Statistical analysis was performed using ANOVA and chi-square test as indicated. P<0.05 (two-tailed) was considered statistically significant.

This study was approved by JBVAMC IRB as expedite protocol (#1455698-1). Written informed consent from patients was not required.

## Results

Characteristics of 166 patients 90 years and older with physician-diagnosed COPD in their health records are shown in Table 1.

**Table 1.**
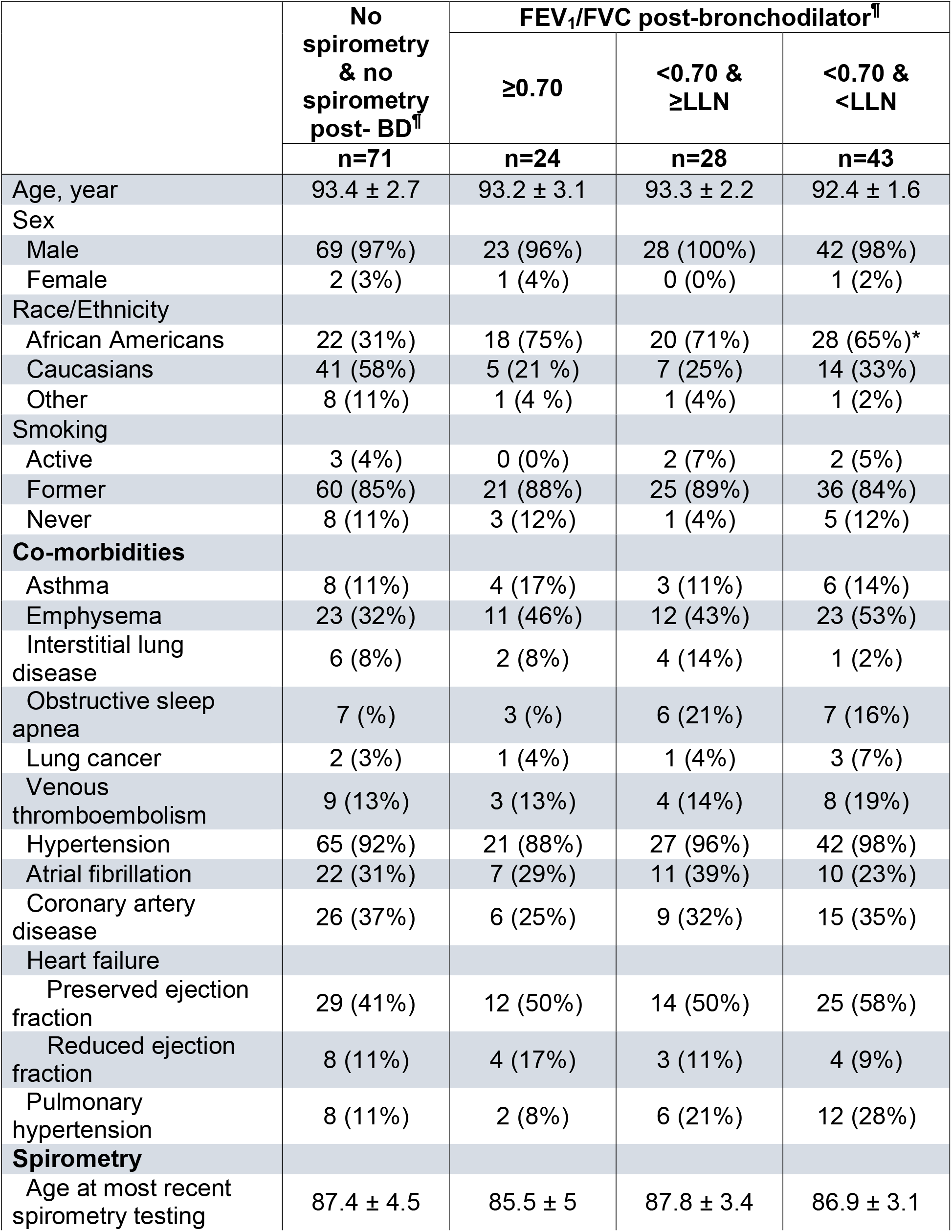
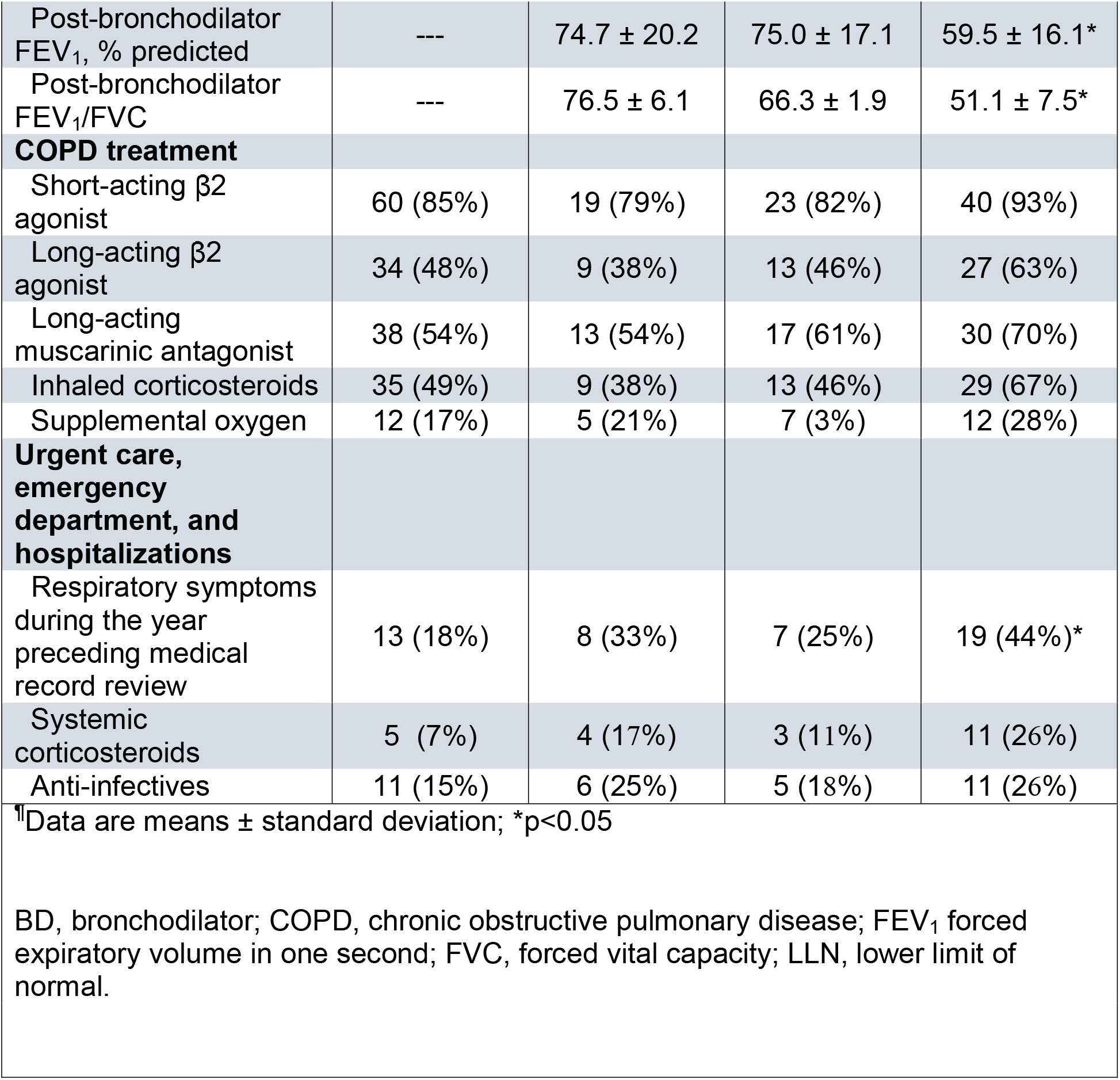
Characteristics of nonagenarians with physician-diagnosed COPD

The patients were predominantly ex-smoker (96%), African American (52%) males (96%). Sixty of 166 patients had no spirometry reports in their health records (36%). In 11 patients (7%), no post-bronchodilator spirometry testing was performed. Patients with adequate spirometry consisted of 1) FEV_1_/FVC≥0.70, n=24; 2) FEV_1_/FVC<0.70 and >LLN, n=28; and 3) FEV_1_/FVC<0.70 and <LLN, n=43 (Table 1). There were no significant differences in age, sex, smoking history, prevalence of co-morbidities, and prescribed COPD medications, including systemic corticosteroids and anti-infectives during urgent care and emergency department visits, between the four groups.

Although only 61 of 166 patients in groups 2&3 (37%) fulfilled GOLD guidelines [21], COPD medications, predominantly short-acting β_2_ agonist and long-acting muscarinic antagonist, were prescribed to 95 patients in groups 1&2 (Table 1). The number of patients with FEV_1_/FVC <0.70 and <LLN post-bronchodilator seen in urgent care and emergency department and hospitalized for respiratory symptoms during the year preceding medical record review (19/43, 44%) was significantly higher than that of patients without spirometry testing or post-bronchodilator spirometry testing (13/71, 18%) (p<0.05) (Table 1). In addition, 9 of 95 (9%) patients with no spirometry, no post-bronchodilator spirometry, or spirometry with post-bronchodilator FEV_1_/FVC >0.70 were treated with systemic corticosteroids during urgent care and emergency department visits (Table 1).

## Discussion/Conclusion

The new finding of this study is that a substantial proportion of nonagenarians at our medical center are overdiagnosed with COPD and treated accordingly. This observation is ominous because COPD medications, such as short- and long-acting bronchodilators, may predispose nonagenarians, particularly those with cardiovascular co-morbidities, to serious adverse events given the altered pharmacokinetics of drugs and polypharmacy with drug-drug interactions in this aging patient population [11, 13-15, 23-27]. To that end, older people are often under-represented and those with co-morbidities excluded from clinical trials of safety and efficacy of long-acting bronchodilators in patients with COPD [11, 16]. Importantly, focusing on COPD management, including inappropriate treatment with systemic corticosteroids during acute care events as observed in this study, may miss other treatable medical conditions and adversely affect patient outcomes. Hence, prescribing COPD medications to nonagenarians should be reserved only for those with COPD diagnosed by current practice guidelines with spirometry performed before and after inhaling a short-acting β_2_-agonist [21].

Diagnosing COPD in nonagenarians is, however, challenging due to age-dependent decline in lung function, presence of comorbidities that could masquerade as COPD, such as heart failure and muscle deconditioning, and cognitive difficulties that may compromise successful implementation of COPD treatments [2, 3, 6-8]. Hankinson et al [4] showed that at age 70 years, the expected FEV_1_/FVC is ∼74%, a value close to the post-bronchodilator 70% cut-off currently used to diagnose COPD. However, the authors did not report the expected FEV_1_/FVC in nonagenarian persons. Nonetheless, these observations highlight the need for using both post-bronchodilator FEV_1_/FVC <0.70 and <LLN to diagnose COPD in nonagenarians. Unfortunately, we found that this approach is not universally adopted by physicians at our institution. Conceivably, underuse of spirometry in nonagenarians could lead to nonspecific symptoms being labeled as COPD without post-bronchodilator spirometry performed to confirm the diagnosis [9, 10, 16]. Accordingly, we propose that health care providers caring for nonagenarians should be reminded about the need for pre- and post-bronchodilator spirometry to diagnose and treat COPD in this patient population.

Our study has a relatively large sample size comprising predominantly of nonagenarian African American males, an underrepresented patient population in the COPD space [5, 6]. Thus, our findings add to the paucity of medical literature on overdiagnosis and treatment of COPD in nonagenarians [2, 3, 6, 10].

Several limitations of this study are notable. It is a retrospective, single center study comprised almost exclusively of males. Hence, generalizability of our observations is limited. Conceivably, spirometry could have been performed in our patients long before the age 90 at a time when electronic health records were not available and paper records of these tests were discarded with passage of time. This, in turn, may have accounted, in part, for substantial number of patients not having spirometry reports in their current electronic health records. To overcome this problem, we propose that spirometry should be performed in symptomatic nonagenarians with physician-diagnosed COPD whenever spirometry reports are not available in medical records. Whether other proposed means to diagnose COPD in symptomatic nonagenarians who cannot adequately perform spirometry, such as CT scan of the chest and compatible history and clinical manifestations of COPD, could be used instead remains to be determined [16, 21].

In summary, these data suggest that a large proportion of nonagenarians at our medical center are overdiagnosed with and treated for COPD. A larger, multi-center prospective study is warranted to support or refute these retrospective observations.

## Data Availability

All data produced in the present work are contained in the manuscript.

## Acknowledgments

The views expressed in this article are those of the authors and do not necessarily reflect the position or policy of the Department of Veterans Affairs or the United States government.

This material is the result of work supported with resources and the use of facilities at the Jesse Brown VA Medical Center, Chicago, Illinois, USA.

## Author Contributions

All co-authors contributed equally to study concept, design, data acquisition, analyses and interpretation, and drafting the manuscript. The final version of the manuscript was reviewed by all co-authors and approved for submission.

## Data Availability Statement

All data generated or analyzed during this study are included in this article [and/or] its supplementary material files. Further enquiries can be directed to the corresponding author.

